# Ancestry-associated immune signalling differences in atopic dermatitis and dupilumab treatment

**DOI:** 10.64898/2025.12.08.25341816

**Authors:** Duan Ni, Ralph Nanan

## Abstract

**Background:** Atopic dermatitis (AD) is a common atopic disease worldwide and dupilumab, a monoclonal antibody directing to the IL4/IL13 signalling, is emerging as an effective therapy for AD. Recently, there is report describing differences in AD severity and treatment responses to dupilumab among different geographic regions, but the underlying mechanisms remain unresolved. Patients’ ancestral backgrounds represent one of the key differences among various geographic areas. Their implications in variability regarding diseases and treatment responses are gaining more and more recognitions.

**Methods:** We aimed to delineate the potential ancestry-associated differences in AD and treatment responses to dupilumab. We thoroughly surveyed Gene Expression Omnibus (GEO) for transcriptomic dataset in the context of AD and dupilumab treatment involving individuals of diverse ancestral backgrounds and carried out comparative analyses for samples from different ancestral groups.

**Results:** Only one transcriptomic dataset was found for biopsy specimens from lesion and non-lesion skin from AD patients of self-reported *White* and *Asian* ancestral backgrounds. Despite comparable clinical phenotypes, Gene Set Enrichment Analysis revealed that skin samples from White AD patients exhibited upregulated IL4 & IL13 signalling from baseline to up to 4-week post dupilumab treatment, relative to Asian ones.

**Conclusions:** This is the first study of its kind to unravel the ancestry-related differences in AD and dupilumab treatment responses. These findings might be instrumental to future clinical patient stratification, risk assessment and guide personalized medicine treatment options for dupilumab.

---

Recent publication by De Vriese *et al*., *Geographic Differences in Dupilumab Treatment Outcomes for Atopic Dermatitis: A Systematic Review and Meta-Regression Analysis*, systematically reviewed atopic dermatitis (AD) manifestation and dupilumab treatment in East Asian, Norther/Centra Europe and Southern Europe. They reported East Asians having the highest baseline AD eczema area severity index (EASI) scores and Southern Europeans exhibiting the greatest EASI reduction upon dupilumab treatment [1]. However, the underlying mechanisms for these observations remain unresolved.

One of the key differences of these geographic regions are the participants’ ancestral backgrounds. Despite continuing to generate debate, self-reported ancestral backgrounds remain an essential variable in biomedical studies. There is also growing evidence suggesting that self-reported ancestral backgrounds are associated with corresponding genetic profiles [2], which are linked to ancestral differences in disease manifestations and treatment responses, as reported by us and others [3, 4].

We thus endeavour to elucidate the potential implications of ancestral backgrounds on AD and dupilumab treatment responses. We hypothesized that data from individuals of diverse ancestral backgrounds might shed light on these issues. For this purpose, we exhaustively surveyed Gene Expression Omnibus (GEO) for transcriptomic dataset in the context of AD and dupilumab treatment involving individuals of diverse ancestral backgrounds. Only one study was found (GEO ID: GSE130588) [5], demonstrating the underrepresentation of ancestral factors in AD research and treatment.

GEO130588 analyzed the lesion and non-lesion skin biopsies from AD patients throughout 16-week dupilumab treatment at a transcriptomic level, based on a multi-centre, placebo-controlled, double-blind clinical trial (NCT1979016). It covered individuals with self-reported ancestral backgrounds of “*White*” (not Hispanic or Latino) versus “*Asian*” (Figure 1A). At baseline week 0 (W0), EASI score and SCOring Atopic Dermatitis (SCORAD) and gender profiles were similar in both ancestral groups (Figure 1B-C). They also exhibited comparable reductions in EASI and SCORAD scores as treatment responses to dupilumab (Figure 1D).

**Figure 1.**
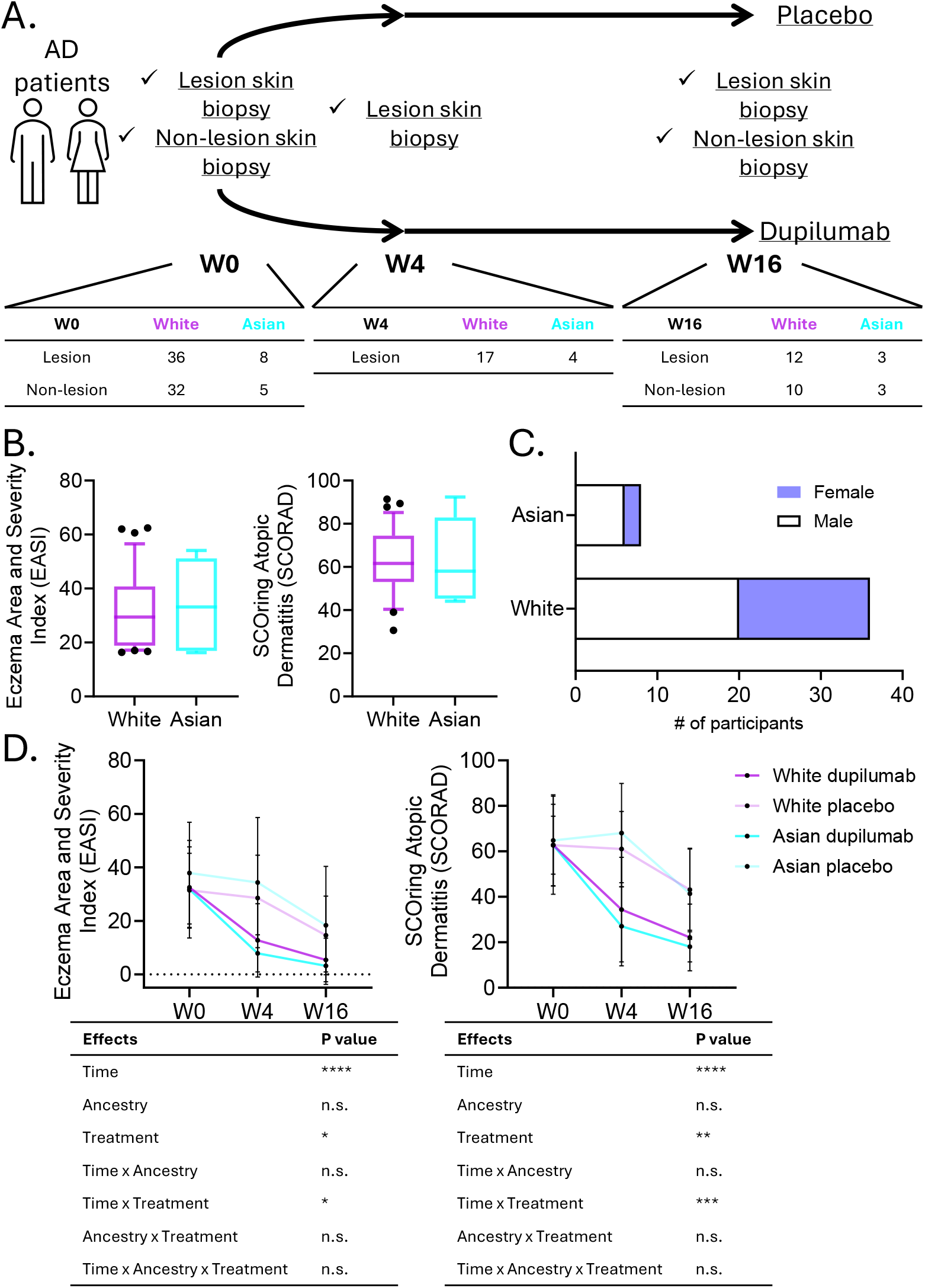
**A**. Schematic overview of the study design of the multi-centre, placebo-controlled, double-blinded trial (NCT01979016) evaluating the efficacy, safety and effects of dupilumab on atopic dermatitis (AD), which the transcriptomic dataset GEO130588 is based on. In brief, AD patients were randomized 1:1 to placebo or weekly subcutaneous doses of 200mg of dupilumab for 16 weeks. Skin biopsy specimens were collected from lesion (baseline Week0, W0; Week4, W4; and Week16, W16) and non-lesion (W0 and W16) skin and subject to transcriptomic analysis. For the purposes of our current study, we focused on participants from self-reported White and Asian ancestral backgrounds and comparatively analzyed their transcriptomic profiles throughout the trial. **B**. Box charts for the baseline (W0) Eczema Area and Severity Index (EASI, left) and SCOring Atopic Dermatitis (SCORAD, right) scores of AD patients from White (purple) versus Asian (cyan) ancestral backgrounds. The boxes extend between 25^th^ to 75^th^ percentiles, while the whiskers mark 10^th^ to 90^th^ percentiles, and the lines in the middle denote the medians. **C**. Bar chart visualizing the gender profiles of AD patients from White versus Asian ancestral backgrounds, with white bars reflecting males and blue bars representing females. χ2 test found no difference in the gender profiles comparing White versus Asian ancestral groups. **D**. Longitudinal changes of EASI and SCORAD scores of AD patients from White (purple) versus Asian (cyan) ancestral backgrounds throughout the trial. Participants receiving placebo or dupilumab are denoted with lighter or darker colours respectively. Three-way ANOVA revealed significant improvement of EASI and SCORAD scores from dupilumab treatment longitudinally, but no difference was found between White and Asian ancestral groups. (*: p<0.05, **: p<0.01, ***: p<0.0005, ****: p<0.0001, n.s.: not significant)

Mechanistically, we performed Gene Set Enrichment Analysis (GSEA) for the transcriptomic data for skin biopsy samples from different timepoints, comparing White and Asian ancestral groups. At W0, 134 and 117 pathways were upregulated in White versus Asian individuals in their lesion and non-lesion skin areas respectively (Figure 2A). Among 34 pathways elevated in participants of White ancestral background at both lesion and non-lesion sites, “IL4&IL13 signalling pathway” (main target of dupilumab), and “IL10 signalling pathway” consistently featured. (Figure 2B). This pattern persisted in lesion skin areas at 4-week post dupilumab treatment (W4, Figure 2C) but dissipated in W4 non-lesion skin and in 16-week post treatment timepoint.

**Figure 2.**
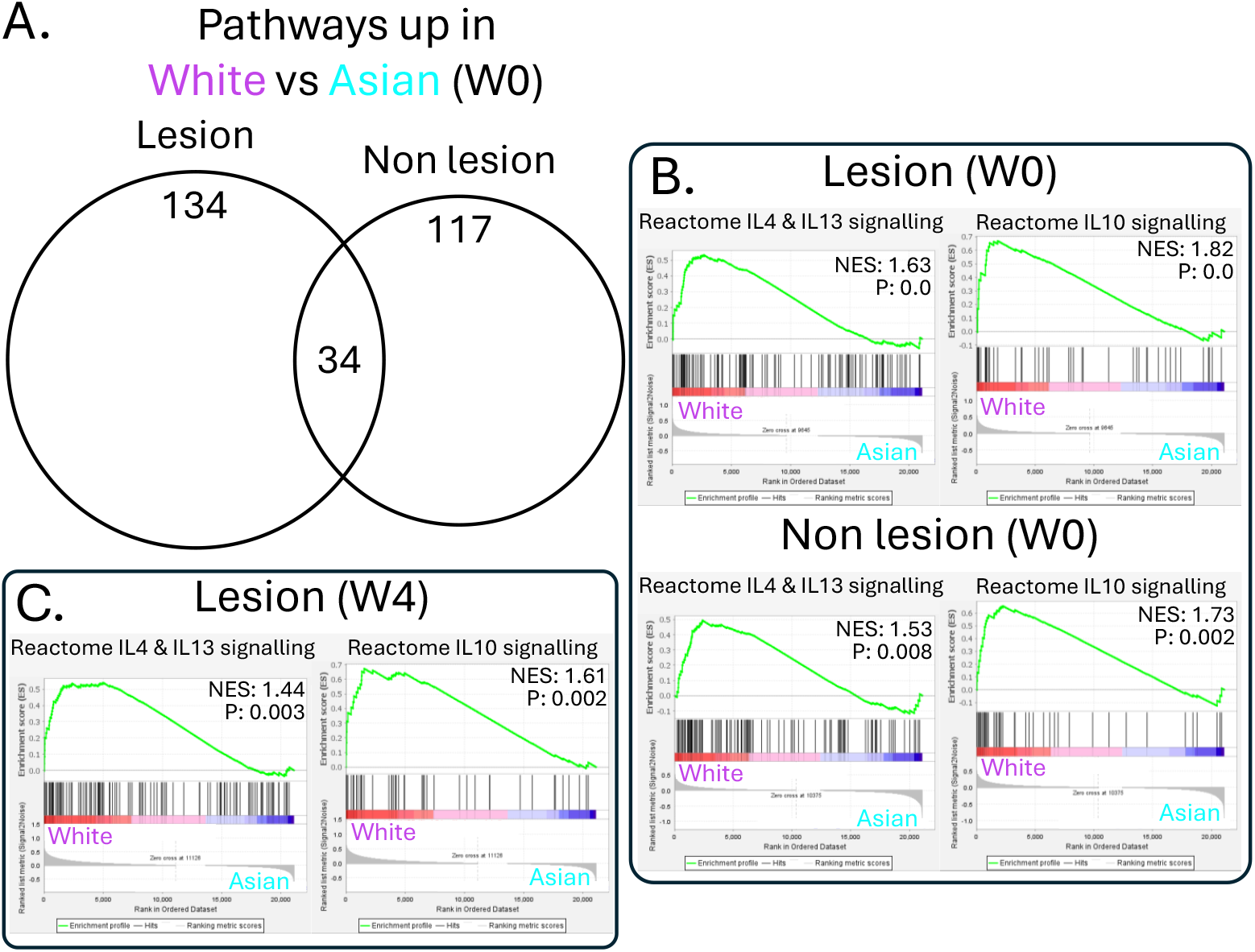
**A**. Skin biopsy transcriptomic data were analyzed by GSEA based on the *Reactome* dataset. At baseline (W0), 134 and 117 pathways were upregulated in White individuals relative to Asian individuals in biopsy specimens from lesion and non-lesion skin respectively. 34 pathways were elevated in White individuals at both sites. **B**. GSEA plots showing increase in IL4 & IL13 signalling (left) and IL10 signalling (right) in samples from White (purple) versus Asian (cyan) individuals in their lesion (upper) and non-lesion (lower) skin at baseline W0. **C**. GSEA plots showing increase in IL4 & IL13 signalling (left) and IL10 signalling (right) in samples from White (purple) versus Asian (cyan) individuals in their lesion skin biopsy at W4 timepoint. (NES: normalized enrichment score; P: p value)

Collectively, this represents to our knowledge the first study investigating the mechanistic differences in AD and dupilumab treatment responses comparing individuals of different ancestral backgrounds. We found that despite similar clinical manifestations, White AD patients displayed higher IL4 and IL13 signalling from baseline to up to 4-week of dupilumab treatment. These findings suggest that ancestral background differences across geographic regions are unlikely to account for the geographic variations in AD and dupilumab responses reported by De Vriese *et al*., although validation in additional cohorts from diverse locations is needed.

Comparisons of different ancestral or geographic backgrounds are inevitably subject to various confounders like environmental and socioeconomic factors. Our analyses based solely on AD cohorts from North America might be better controlled compared to the original systematic review looking into studies across different continents. Notwithstanding these factors, the ancestral differences we found might have implications in informing AD patient clinical stratifications and personalized medicine treatment options with dupilumab regarding dosage and duration. Ultimately, our findings also highlight the pressing need to increase ancestral diversity in atopic research, promoting health equity.

## Data Availability

Data used in this study is available at: https://www.ncbi.nlm.nih.gov/geo/query/acc.cgi?acc=GSE130588

## Author contributions

Study conceptualization and design: D.N. R.N. Investigation and data curation: D.N. Data analysis: D.N. Funding and resources: R.N. Manuscript preparation: D.N., R.N. All authors read, edited and approved the final version of the manuscript.

## Acknowledgements

D.N. and R.N. were supported by the Norman Ernest Bequest Fund.

## Ethics statement

Not applicable.

