## Supplementary Information for "Ancestry-associated immune signalling differences in atopic dermatitis and dupilumab treatment"

### Materials and Methods

*Data collection and analysis*

We aim to comparatively analyze the transcriptomic profiles of atopic dermatitis (AD) patients of different ancestral backgrounds and their responses to dupilumab treatment. Therefore, we exhaustively survey Gene Expression Omnibus (GEO) for any transcriptomic data in these regards. Most prior studies did not disclose the ancestral backgrounds of their participants. Only one dataset was found with detailed ancestral background information (GSE130588), which stemmed from a placebo-controlled, double-blind trial (NCT01979016) examining the efficacy, safety, and effect of dupilumab in treating AD patients [1], covering individuals from both self-reported *White* versus *Asian* ancestral backgrounds.

Demographic background information and clinical data were obtained from the original clinical trial [1], and the corresponding processed transcriptomic data were downloaded from GEO (GSE130588). The data were then analyzed with Gene Set Enrichment Analysis (GSEA) following the software's tutorial based on the *Reactome* dataset [2], comparing samples of White versus Asian ancestral backgrounds. Gene sets with differences of p value smaller than 0.05 were considered to be significant.
